# Anti-vaccine attitudes and risk factors for not agreeing to vaccination against COVID-19 amongst 32,361 UK adults: Implications for public health communications

**DOI:** 10.1101/2020.10.21.20216218

**Authors:** Elise Paul, Andrew Steptoe, Daisy Fancourt

## Abstract

**Background:** Negative attitudes towards vaccines and an uncertainty or unwillingness to receive vaccinations are major barriers to managing the COVID-19 pandemic in the long-term. We estimate predictors of four domains of negative attitudes towards vaccines and identify groups most at risk of uncertainty and unwillingness to receive a COVID-19 vaccine in a large sample of UK adults.

**Methods:** Data were from 32,361 adults in the UCL COVID-19 Social Study. Ordinary least squares regression analyses examined the impact of socio-demographic and COVID-19 related factors on four types of negative vaccine attitudes: mistrust of vaccine benefit, worries about unforeseen effects, concerns about commercial profiteering, and preference for natural immunity. Multinomial logistic regression examined the impact of socio-demographic and COVID-19 related factors, negative vaccine attitudes, and prior vaccine behaviour on uncertainty and unwillingness to be vaccinated for COVID-19.

**Findings:** 16% of respondents displayed high levels of mistrust or misinformation about vaccines across one or more domains. Distrustful attitudes towards vaccination were higher amongst individuals from ethnic minority backgrounds, with lower levels of education, lower annual income, poor knowledge of COVID-19, and poor compliance with government COVID-19 guidelines. Overall, 14% of respondents reported unwillingness to receive a vaccine for COVID-19, whilst 22% were unsure. The largest predictors of both COVID-19 vaccine uncertainty and refusal were low income (< £30,000 a year), having not received a flu vaccine last year, poor adherence to COVID-19 government guidelines, female gender, and living with children. Amongst vaccine attitudes, intermediate to high levels of vaccine benefit mistrust and concerns about future unforeseen side effects were the most important determinants of both uncertainty and unwillingness to vaccinate against COVID-19.

**Interpretation:** Negative attitudes towards vaccines are major public health concerns in the UK. General mistrust in vaccines and concerns about future side effects in particular will be barriers to achieving population immunity to COVID-19 through vaccination. Public health messaging should be tailored to address these concerns.

**Funding:** The Nuffield Foundation [WEL/FR-000022583], the MARCH Mental Health Network funded by the Cross-Disciplinary Mental Health Network Plus initiative supported by UK Research and Innovation [ES/S002588/1], and the Wellcome Trust [221400/Z/20/Z and 205407/Z/16/Z].

Evidence before this study
We searched PubMed for articles published in English from 1 January 2020 to 20 September 2020 with the following keywords: (“COVID19 vaccine” OR “coronavirus vaccine”) and (“intent*” OR “refusal”). Our search found 639 titles. Several previous studies have examined predictors of intent to vaccinate for COVID-19 when it becomes available. Reasons for unwillingness to receive the COVID-19 vaccination when it becomes available centred on concerns about its newness, safety, and potential side effects. However, estimates and predictors of negative vaccine attitudes in general and how these attitudes predict uncertainty and unwillingness to vaccinate in the context of COVID-19 are unavailable.

Added value of this study
The attitudinal and behavioural barriers to being unsure about receiving a COVID-19 vaccine and not intending to receive one were largely overlapping; 1) didn’t get a flu vaccine last year, 2) poor adherence to government guidelines, 3) concerns about the unforeseen future effects of vaccines, and 4) and general mistrust in the benefits of vaccines.

Implications of all of the available evidence
Mistrust towards vaccines represent a significant challenge in achieving the vaccination coverage required for population immunity. Taken together, there is evidence that groups most vulnerable to falling ill and dying of COVID-19 (e.g. those from ethnic minority backgrounds and who have lower incomes) have more negative attitudes towards vaccines and are less willing to vaccinate against COVID-19. Not everyone who intends to receive a COVID-19 vaccination will be able to do so because of practical barriers such as lack of accessibility and government decisions on the availability of the vaccine, underscoring the importance of improving vaccine attitudes in the general population to improve vaccine uptake amongst those who are offered a vaccine and prevent widening socio-economic health inequalities. Vaccine safety communication to increase public trust by the time a COVID-19 vaccine is available should begin now.

## Introduction

The long-term success of the public health response to the coronavirus disease 2019 (COVID-19) pandemic will depend on acquired immunity in a sufficient proportion of the population (herd immunity), which is estimated to be 67% for COVID-19.^1^ Achieving population immunity through natural means, or by allowing a large proportion of the population to become infected, would cause unprecedented strain on healthcare resources and could result in up to 30 million deaths worldwide.^1^ Widespread vaccination is therefore essential for managing COVID-19 transmission.^2^ However, the current pandemic is occurring amidst a backdrop of widespread mistrust in the safety and effectiveness of vaccines globally.^3^ Thousands of people have taken to the streets around the world to protest COVID-19 social distancing policies and the prospect of mass vaccinations. This is concerning as public attitudes towards vaccine safety, their importance, and effectiveness are consistently associated with vaccine uptake.^3^ Although general population data from the UK and Europe indicate mostly positive attitudes towards vaccines, research is suggesting there is still a substantial (≅ 10%) proportion of adults who are unsure of or distrust the safety and effectiveness of vaccines in the UK and Europe general population.^4^

Findings from nationally representative studies suggest unwillingness and uncertainty about receiving a COVID-19 vaccine will be a significant challenge in achieving the vaccination coverage required for population immunity. Early in the pandemic (April 2020), 26% of adults across seven European countries including the UK were unsure or unwilling to get a COVID-19 vaccine when available.^5^ Other studies have found that around one-quarter of French^6^ and US^7^ adults do not intend to receive the vaccine even if offered it. Research conducted later in the pandemic, in mid-July after restrictions had started to ease, revealed that an even greater proportion of the UK adult population (36%) was either unsure or definitely would not get the vaccine.^8^ Women,^5,6,9,10^ those with lower levels of education,^6,8,10^, low income,^6,7,10^ and who were not vaccinated against the flu in the past year are more likely to say they will refuse a COVID-19 vaccine when it becomes available.^8,11^

Concerns identified to date for intending not to receive the COVID-19 vaccine include worries about the newness and safety of the vaccine as well as about potential side effects.^5,8,10,12^ The only study that has examined associations between general vaccine attitudes and intent to vaccinate against COVID-19 found confidence in vaccine safety to be the largest determinant.^7^ However, to our knowledge no study has examined predictors of vaccine attitudes and how these attitudes in turn relate to an unwillingness to vaccinate in the context of the COVID-19 pandemic. Further missing from this work is information on determinants of uncertainty about receiving the COVID-19 vaccine, as prior research has only examined vaccine intent outcomes as binary (e.g. willing vs. unwilling)^5,6,9^ or as a continuous measure of vaccine likelihood.^8^ Understanding factors driving uncertainty about being vaccinated against COVID-19 is crucial, as individuals who are uncertain may be the most realistic targets for public health communications programmes encouraging vaccination.^13^ As these individuals make up a greater share of the population than those who are certain they would not vaccinate, understanding their concerns is paramount.^5,8,9^

Consequently, there is an urgent need for a more updated and nuanced understanding of attitudes towards vaccines and factors determining vaccine intent in the context of the COVID-19 pandemic in order to tailor public health messaging accordingly.^14^ Therefore, the aims of the present study were to identify factors predictive of (1) a range of negative attitudes towards vaccines, and (2) uncertainty and lack of intent to vaccinate against COVID-19. Importantly, we utilise a large sample of UK adults who were asked about their vaccine attitudes and intentions at the beginning of a second wave of the COVID-19 pandemic (September 2020).^15^ Exploring predictors of vaccine attitudes in general terms has the potential to help policymakers identify and adapt interventions that increase vaccine confidence that have previously been tested outside the COVID-19 pandemic. Findings have public health importance for the design of interventions aimed at maximising uptake of the COVID-19 vaccine among the general population.

## Methods

### Study design and participants

Data were drawn from the COVID-19 Social Study; a large panel study of the psychological and social experiences of over 75,000 adults (aged 18+) in the UK during the COVID-19 pandemic. The study commenced on 21 March 2020 and involves online weekly data collection from participants for the duration of the COVID-19 pandemic in the UK. The study is not random and therefore is not representative of the UK population. But it does contain a well-stratified sample that was recruited using three primary approaches. First, snowballing was used, including promoting the study through existing networks and mailing lists (including large databases of adults who had previously consented to be involved in health research across the UK), print and digital media coverage, and social media. Second, more targeted recruitment was undertaken focusing on (i) individuals from a low-income background, (ii) individuals with no or few educational qualifications, and (iii) individuals who were unemployed. Third, the study was promoted via partnerships with third sector organisations to vulnerable groups, including adults with pre-existing mental health conditions, older adults, carers, and people experiencing domestic violence or abuse. The study was approved by the UCL Research Ethics Committee [12467/005] and all participants gave informed consent.

For these analyses, we focused on individuals gave data during the month in which the vaccine module was administered (7 September to 5 October 2020). A total of 32,585 participants met this criterion. We then excluded participants with any missing data on vaccine outcome variables (*n* = 12). Due to insufficient statistical power, we also excluded individuals who had selected “other” in response to gender (*n* = 134) and “prefer not to say” on ethnicity (*n* = 95). Seventeen of these individuals selected both responses, leaving a total analytical sample size of 32,361.

## Measures

### Outcome variables

Negative general attitudes towards vaccines were measured using the 12-item Vaccination Attitudes Examination (VAX) Scale. Participants were asked to focus on vaccines in general rather than specifically on vaccines for COVID-19. Responses were rated on a 6-point scale from 1 “strongly agree” to 6 “strongly disagree.” Four subscales which have previously been derived^16^ were calculated: 1) mistrust of vaccine benefit, (2) worries about unforeseen future effects, (3) concerns about commercial profiteering, and (4) preference for natural immunity. Adequate convergent validity and internal reliability was established for all 4 subscales in two adult samples (Cronbach’s alphas = 0·77-0·93). Internal consistencies in the current sample were good (Cronbach’s alphas 0·91-0·94). Each of the four subscales was grouped into high (a score of 5-6 on a scale of 1-6), intermediate (score of 3-4), and low (score of 1-2) levels of negative attitudes towards vaccines.

Uncertainty and unwillingness to vaccinate against COVID-19 when available were based on one item (see Supplemental Table S1 for question wording). Response options ranged from “1- very unlikely” to “6 – very likely”. An ordinal variable was coded: (0) intend to vaccinate (responses of 5-6), (1) unsure about whether to vaccinate (responses of 3-4), and (2) unwilling to vaccinate (responses of 1-2).

### Predictor variables

Socio-demographic factors included gender (male vs. female), age group (65+, 50-64, 30-45, and 18-29) ethnicity (white vs. BAME groups [i.e. Asian/Asian British, Black/Black British, White and Black/Black British, Mixed race, Chinese/Chinese British, Middle Eastern/Middle Eastern British, or other ethnic group]), education (postgraduate degree, undergraduate degree [further education after the age of 18], A-levels (equivalent to education to age 18) or vocational training, GCSE or lower [equivalent to education to age 16] and no formal qualifications), income (annual household income: >£90,0000, £60,000-89,999,£30,000-59,999, £16,000-29,000, and < £16,000) employment status (not employed vs. employed at the start of the pandemic), living arrangement (live alone vs. not alone), area of dwelling (urban [city, large town, small town] vs. rural [village, hamlet, isolated dwelling]), the presence of a child in the household (no children in the household vs. children in the household), and government’s identified key worker status (not a key worker vs. key worker). The latter included people with jobs deemed essential during the pandemic (e.g. health and social care, education and childcare) and who were required to leave home to carry out this work during the lockdown.

Participant reports of whether they had received clinical diagnoses of a mental illness (e.g. depression, anxiety, or other psychiatric diagnosis) or chronic physical health condition (e.g. high blood pressure, diabetes, heart disease, lung disease (asthma or COPD), cancer, or other physical health condition) were used to create two binary variables (yes/no) to indicate the presence or absence of pre-existing physical and mental health conditions.

Coronavirus anxiety during the past two weeks was measured using the Coronavirus Anxiety Scale (CAS).^17^ The CAS contains 5 items with 5-point responses ranging from “not at all” to “nearly every day”. The scale has shown good internal reliability (Cronbach’s alpha = 0·93), construct validity, diagnostic viability, and equivalency of measurement across demographic groups.^17^ A CAS score of 9 or more classified adults as meeting (90% sensitivity) or not meeting (85% specificity) the threshold for Generalised Anxiety Disorder.^17^ We categorised responses such that participants with one or more COVID-19 anxiety symptom were compared to those who did not report any such symptoms.

Confidence in government and the health service to handle the pandemic were assessed with one question each. Response options ranged from 1 (none at all) to 7 (lots). Two binary variables were created to compare individuals who had a lot of (5-7) versus low (1-4) confidence in the government and health system.

Responses to the question on compliance with government COVID-19 guidelines were on a scale from 1 (none at all) to 7 (very much so). We analyse this as a binary variable reflecting higher (6-7) vs lower (1-5) compliance. Knowledge of COVID-19 was measured with the question: rated on a 7-point scale from 1 (very poor knowledge) to 7 (very good knowledge). Responses of 1-4 were categorised as very poor/poor compared to very good/good (5-7) COVID-19 knowledge. The presence or absence of having had COVID-19 was categorised as a binary variable (yes, diagnosed and recovered, or yes, diagnosed and still ill, or not formally diagnosed but suspected, versus no, not that I know of or no). Prior vaccine behaviour was based on two yes/no questions. See Supplemental Table S1 for a listing of all question wording.

### Statistical analysis

Ordinary least squares (OLS) regressions were carried out to examine socio-demographic and COVID-19- related predictors of each of the four negative attitudes toward vaccines subscales. We then fitted one multinomial logistic regression model to examine associations of socio-demographic and COVID-19- related factors, negative vaccine attitudes, and prior vaccine behaviours with intent to vaccinate against COVID-19. The outcome variable in the latter model was coded such that those likely to vaccinate were compared to i) uncertainty about whether to vaccinate and ii) unwillingness to vaccinate. Results are presented as relative risk ratios (RRR) with corresponding 95% confidence intervals (CI). Missing data

The pattern of missing data in the study sample is presented in Supplemental Table S2. The proportion of missing data ranged from 0.01% for having had COVID-19 to 19.66% for the Coronavirus Anxiety Scale. Multiple imputation by chained equations^18^ was used to generate 50 imputed datasets for each variable in participants with complete data on all vaccine outcomes (N = 32,361). Imputation models included all variables used in the analysis, as well as additional auxiliary variables (home ownership, anxiety symptoms, depressive symptoms, smoking status). Substantives results using cases without any missing data and the imputed sample were similar (Supplemental Tables S3 and S4). To account for the non-random nature of the sample, all data were weighted to the proportions of gender, age, ethnicity and education obtained from the Office for National Statistics.^19^ Analyses were conducted using Stata version 16.^20^

## Results

Characteristics of both the unweighted and weighted samples are presented in Table 1. 7·2% of the sample expressed mistrust of vaccines (e.g. a score of 5-6 on a scale of 1 to 6), whilst 17·2% were uncertain about their levels of trust (a score of 3-4 out of 6). 16·3% expressed strong worries about unforeseen effects, whilst 52·9% expressed moderate worries. 8·1% expressed strong concerns and 28·8% moderate concerns about commercial profiteering. 8·5% expressed a strong preference for natural immunity, whilst 44·7% also expressed some feelings that natural immunity might be better than a vaccine. Correlations among the negative vaccine attitudes scales were medium (mistrust and unforeseen effects: *r* = 0·38, *p* < 0·001; mistrust and preference for natural immunity: *r* = 0·48, *p* < 0·001; mistrust and preference for natural immunity: *r* = 0·48, *p* < 0·001) to large (mistrust and commercial profiteering concerns: *r* = 0·62, *p* < 0·001; unforeseen effects and commercial profiteering concerns: *r* = 0·55, *p* < 0·001; commercial profiteering concerns and preference for natural immunity: *r* = 0·64, *p* < 0·001). 64% of the sample said they intended to receive the COVID-19 vaccine if and when one becomes available, compared with 22% who were uncertain and 14% who were unwilling.

**Table 1.**
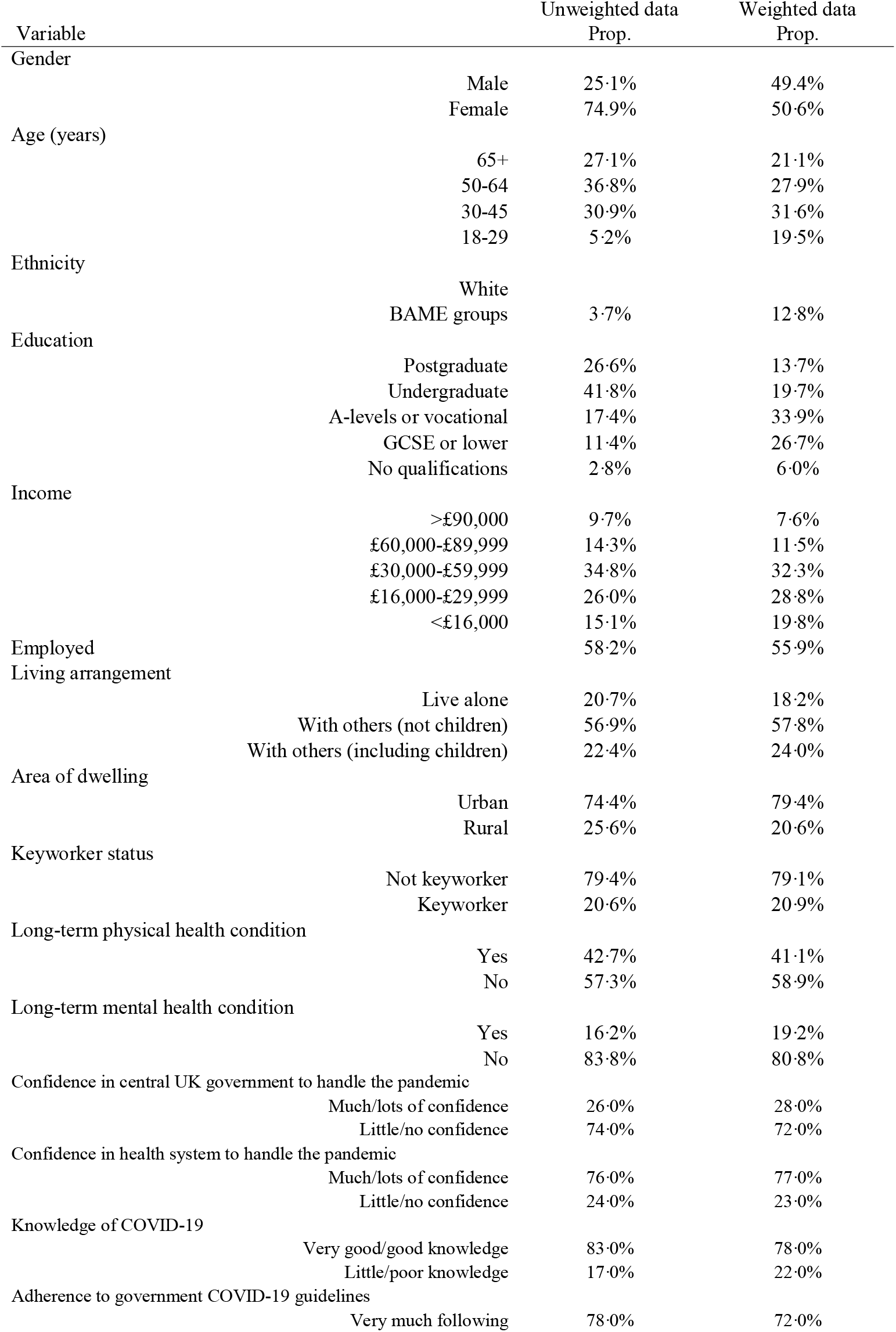
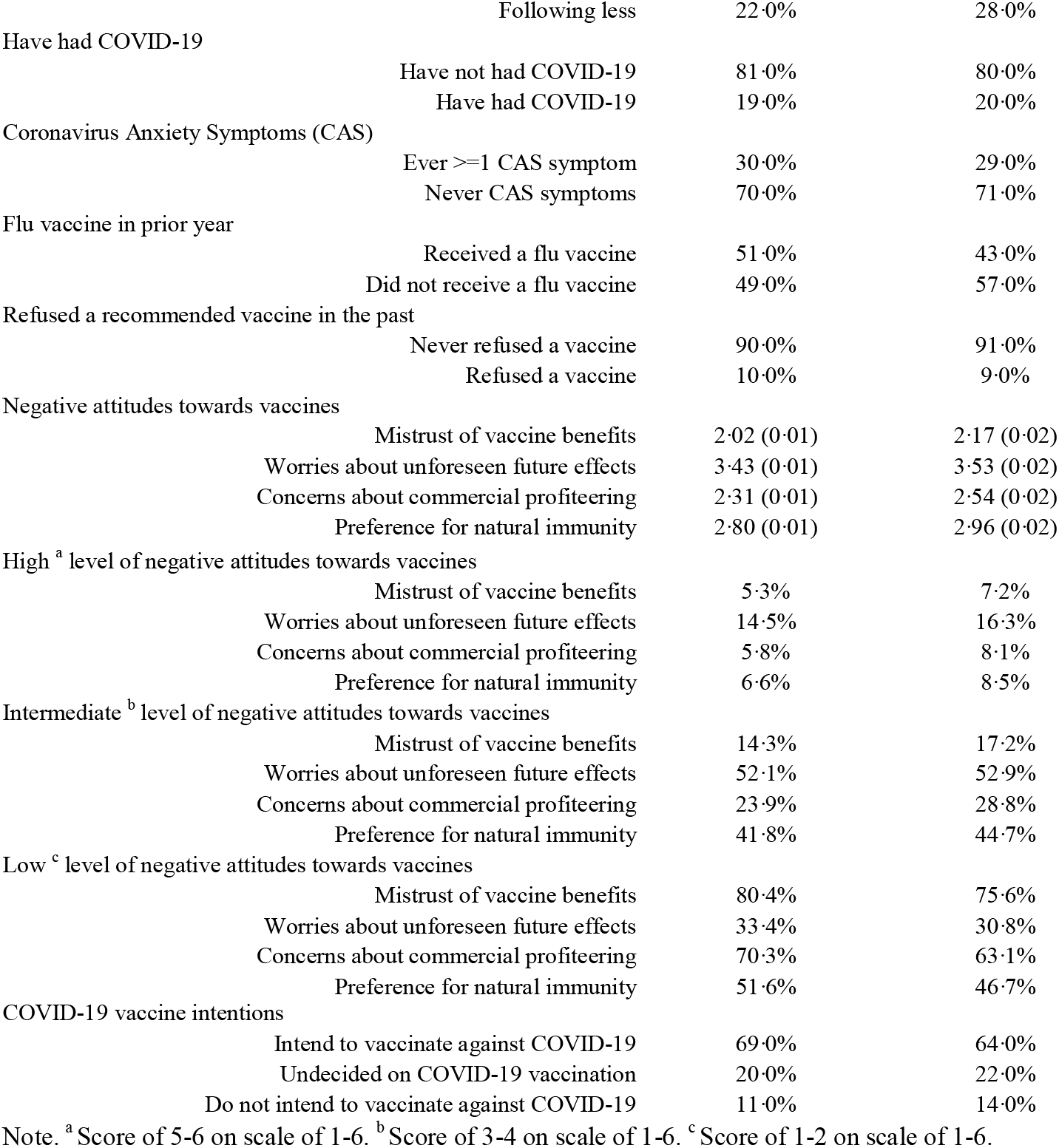
Unweighted and weighted sample characteristics (*N* = 32,361)

| Variable | Unweighted data<br>Prop. | Weighted data<br>Prop. |
| --- | --- | --- |
| Gender |  |  |
| Male | 25.1% | 49.4% |
| Female | 74.9% | 50.6% |
| Age (years) |  |  |
| 65+ | 27.1% | 21.1% |
| 50-64 | 36.8% | 27.9% |
| 30-45 | 30.9% | 31.6% |
| 18-29 | 5.2% | 19.5% |
| Ethnicity |  |  |
| White |  |  |
| BAME groups | 3.7% | 12.8% |
| Education |  |  |
| Postgraduate | 26.6% | 13.7% |
| Undergraduate | 41.8% | 19.7% |
| A-levels or vocational | 17.4% | 33.9% |
| GCSE or lower | 11.4% | 26.7% |
| No qualifications | 2.8% | 6.0% |
| Income |  |  |
| >£90,000 | 9.7% | 7.6% |
| £60,000-£89,999 | 14.3% | 11.5% |
| £30,000-£59,999 | 34.8% | 32.3% |
| £16,000-£29,999 | 26.0% | 28.8% |
| <£16,000 | 15.1% | 19.8% |
| Employed | 58.2% | 55.9% |
| Living arrangement |  |  |
| Live alone | 20.7% | 18.2% |
| With others (not children) | 56.9% | 57.8% |
| With others (including children) | 22.4% | 24.0% |
| Area of dwelling |  |  |
| Urban | 74.4% | 79.4% |
| Rural | 25.6% | 20.6% |
| Keyworker status |  |  |
| Not keyworker | 79.4% | 79.1% |
| Keyworker | 20.6% | 20.9% |
| Long-term physical health condition |  |  |
| Yes | 42.7% | 41.1% |
| No | 57.3% | 58.9% |
| Long-term mental health condition |  |  |
| Yes | 16.2% | 19.2% |
| No | 83.8% | 80.8% |
| Confidence in central UK government to handle the pandemic |  |  |
| Much/lots of confidence | 26.0% | 28.0% |
| Little/no confidence | 74.0% | 72.0% |
| Confidence in health system to handle the pandemic |  |  |
| Much/lots of confidence | 76.0% | 77.0% |
| Little/no confidence | 24.0% | 23.0% |
| Knowledge of COVID-19 |  |  |
| Very good/good knowledge | 83.0% | 78.0% |
| Little/poor knowledge | 17.0% | 22.0% |
| Adherence to government COVID-19 guidelines |  |  |
| Very much following | 78.0% | 72.0% |

|  |  |  |  |
| --- | --- | --- | --- |
|  | Following less | 22.0% | 28.0% |
| Have had COVID-19 |  |  |  |
|  | Have not had COVID-19 | 81.0% | 80.0% |
|  | Have had COVID-19 | 19.0% | 20.0% |
| Coronavirus Anxiety Symptoms (CAS) |  |  |  |
|  | Ever ≥1 CAS symptom | 30.0% | 29.0% |
|  | Never CAS symptoms | 70.0% | 71.0% |
| Flu vaccine in prior year |  |  |  |
|  | Received a flu vaccine | 51.0% | 43.0% |
|  | Did not receive a flu vaccine | 49.0% | 57.0% |
| Refused a recommended vaccine in the past |  |  |  |
|  | Never refused a vaccine | 90.0% | 91.0% |
|  | Refused a vaccine | 10.0% | 9.0% |
| Negative attitudes towards vaccines |  |  |  |
|  | Mistrust of vaccine benefits | 2.02 (0.01) | 2.17 (0.02) |
|  | Worries about unforeseen future effects | 3.43 (0.01) | 3.53 (0.02) |
|  | Concerns about commercial profiteering | 2.31 (0.01) | 2.54 (0.02) |
|  | Preference for natural immunity | 2.80 (0.01) | 2.96 (0.02) |
| High <sup>a</sup> level of negative attitudes towards vaccines |  |  |  |
|  | Mistrust of vaccine benefits | 5.3% | 7.2% |
|  | Worries about unforeseen future effects | 14.5% | 16.3% |
|  | Concerns about commercial profiteering | 5.8% | 8.1% |
|  | Preference for natural immunity | 6.6% | 8.5% |
| Intermediate <sup>b</sup> level of negative attitudes towards vaccines |  |  |  |
|  | Mistrust of vaccine benefits | 14.3% | 17.2% |
|  | Worries about unforeseen future effects | 52.1% | 52.9% |
|  | Concerns about commercial profiteering | 23.9% | 28.8% |
|  | Preference for natural immunity | 41.8% | 44.7% |
| Low <sup>c</sup> level of negative attitudes towards vaccines |  |  |  |
|  | Mistrust of vaccine benefits | 80.4% | 75.6% |
|  | Worries about unforeseen future effects | 33.4% | 30.8% |
|  | Concerns about commercial profiteering | 70.3% | 63.1% |
|  | Preference for natural immunity | 51.6% | 46.7% |
| COVID-19 vaccine intentions |  |  |  |
|  | Intend to vaccinate against COVID-19 | 69.0% | 64.0% |
|  | Undecided on COVID-19 vaccination | 20.0% | 22.0% |
|  | Do not intend to vaccinate against COVID-19 | 11.0% | 14.0% |

Note. <sup>a</sup> Score of 5-6 on scale of 1-6. <sup>b</sup> Score of 3-4 on scale of 1-6. <sup>c</sup> Score of 1-2 on scale of 1-6.

### Predictors of negative attitudes towards vaccines

Results from ordinary least squares regressions predicting four domains of negative vaccine attitudes are presented in Table 2. The strongest associations with negative vaccine attitudes were for variables representing socio-economic status and suggest a gradient such that lower levels of household income and education were associated with progressively more negative views on vaccines across all four domains. In addition, people from BAME groups, those who reported poor compliance with government COVID-19 precautions, and who had low self-rated COVID-19 knowledge also had more negative vaccine views on all four subscales. Women were more likely to express concerns specifically about unforeseen effects of vaccines and less of a preference for natural immunity, as were people without long-term mental health conditions. Low confidence in the health system to handle the pandemic was also associated with greater mistrust of vaccine safety, more worries about unforeseen vaccine effects, and greater concerns about commercial profiteering, whilst low confidence in government to handle the pandemic was associated with lower scores on worries about unforeseen effects and preference for natural immunity. Finally, there was a relationship between previous experience of COVID-19 symptoms and greater negative attitudes. Young people (ages 18-29) were significantly less likely than older adults (ages 65+) to have negative attitudes towards vaccines on all four domains.

**Table 2.**
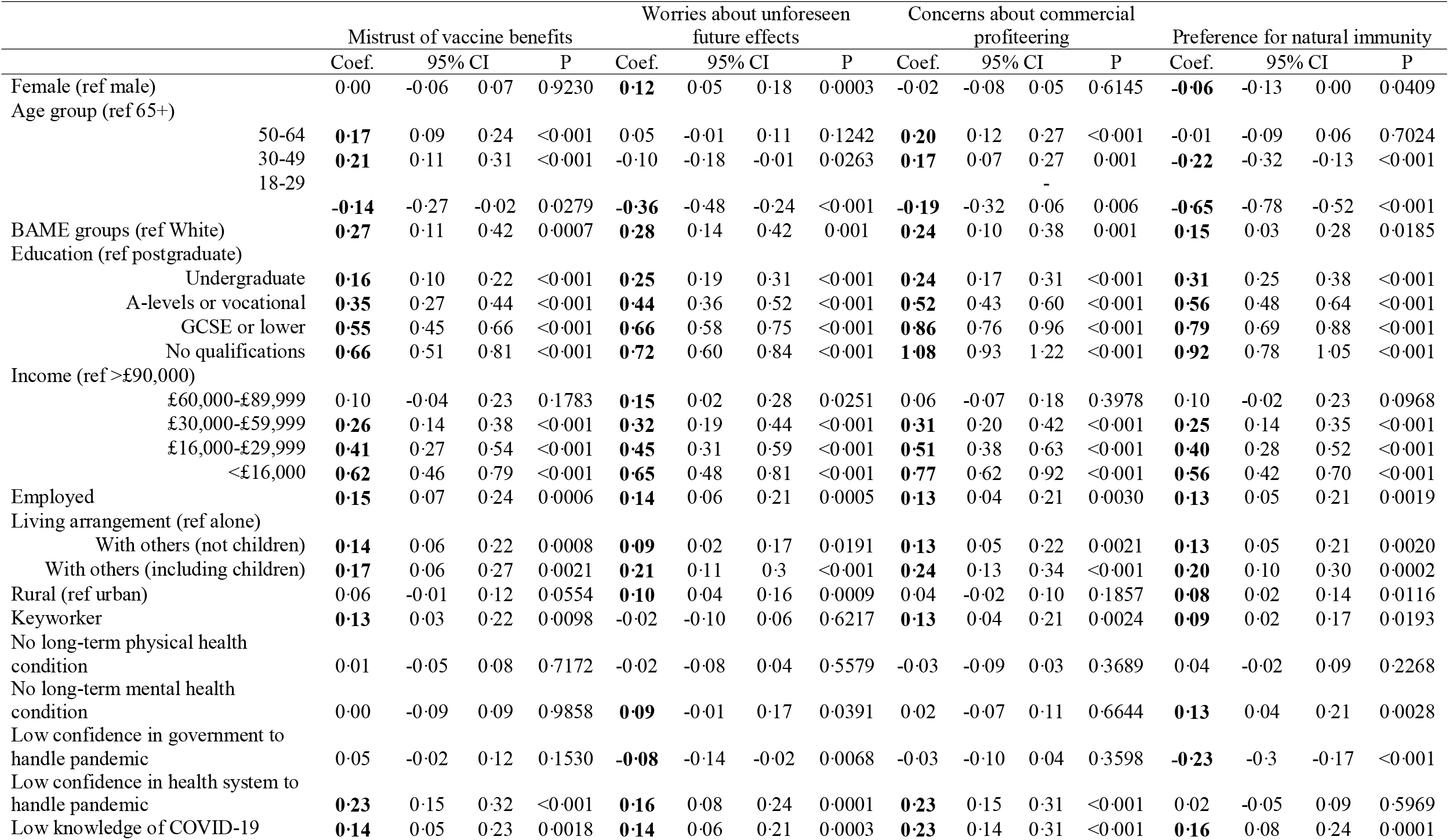
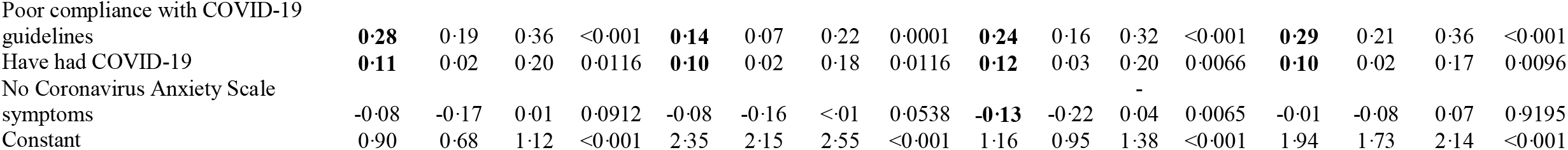
Multivariate linear regression predictors of negative attitudes towards vaccines (weighted, N = 32, 361)

### Predictors of uncertainty and unwillingness to vaccinate against COVID-19

Results from the multinomial logistic regression model predicting risk for uncertainty and a lack of intent to vaccinate against COVID-19 are shown in Table 3. Misinformation and mistrust in vaccines across all four domains (most strongly concerns about unforeseen side effects and mistrust in the benefit of vaccines) were associated with a greater likelihood of vaccination uncertainty and unwillingness for COVID-19. Strong and intermediate levels of mistrust of vaccine benefits were each associated with a 5 times higher relative risk of being unwilling to get a COVID-19 vaccine. Similarly, vaccine unwillingness was predicted by strong worries about unforeseen effects (RRR = 4·91; 95% CI: 3·76 to 6·42), intermediate (but not strong) concerns about commercial profiteering (RRR = 1·73; 95% CI: 1·34 to 2·24), and strong preference for natural immunity (RRR = 2·51; 95% CI: 1·78 to 3·53).

**Table 3.** Predictors of uncertainty and unwillingness to vaccinate against COVID-19 using multivariate multinomial logistic regressions (weighted, *N* = 32,361)

|  | Undecided |  |  |  | Unwilling |  |  |  |
| --- | --- | --- | --- | --- | --- | --- | --- | --- |
|  | RRR | 95% CI |  | P | RRR | 95% CI |  | P |
| Female (ref male) | **1.45** | 1.27 | 1.65 | <0.001 | **1.52** | 1.24 | 1.86 | <0.001 |
| Age (ref 65+) |  |  |  |  |  |  |  |  |
| 50-64 | 1.09 | 0.92 | 1.30 | 0.306 | **1.33** | 1.04 | 1.68 | 0.020 |
| 30-49 | 1.16 | 0.93 | 1.43 | 0.185 | **1.56** | 1.16 | 2.11 | 0.004 |
| 18-29 | 1.04 | 0.79 | 1.38 | 0.775 | 1.23 | 0.76 | 2.00 | 0.405 |
| BAME groups (ref White) | 1.23 | 0.97 | 1.56 | 0.091 | 1.34 | 0.93 | 1.94 | 0.115 |
| Education (ref postgraduate) |  |  |  |  |  |  |  |  |
| Undergraduate | 0.95 | 0.82 | 1.09 | 0.451 | **1.27** | 1.02 | 1.57 | 0.029 |
| A-levels or vocational | 1.08 | 0.90 | 1.29 | 0.428 | **1.74** | 1.35 | 2.24 | <0.001 |
| GCSE or lower | 1.06 | 0.88 | 1.28 | 0.563 | **2.24** | 1.68 | 2.99 | <0.001 |
| No qualifications | 1.10 | 0.82 | 1.47 | 0.516 | **2.33** | 1.56 | 3.47 | <0.001 |
| Income (ref >£90,000) |  |  |  |  |  |  |  |  |
| £60,000-£89,999 | 1.07 | 0.82 | 1.41 | 0.621 | 0.97 | 0.62 | 1.53 | 0.894 |
| £30,000-£59,999 | **1.37** | 1.08 | 1.74 | 0.009 | 1.31 | 0.89 | 1.92 | 0.169 |
| £16,000-£29,999 | **1.47** | 1.13 | 1.90 | 0.004 | **1.59** | 1.06 | 2.41 | 0.026 |
| <£16,000 | **2.10** | 1.56 | 2.82 | <0.001 | **2.16** | 1.37 | 3.41 | 0.001 |
| Employed | **1.24** | 1.05 | 1.47 | 0.012 | 1.24 | 0.97 | 1.58 | 0.082 |
| Living arrangement (ref alone) |  |  |  |  |  |  |  |  |
| With others (not children) | 1.11 | 0.94 | 1.32 | 0.209 | 1.17 | 0.94 | 1.45 | 0.162 |
| With others (including children) | **1.38** | 1.13 | 1.70 | 0.002 | **1.60** | 1.24 | 2.08 | <0.001 |
| Rural (ref urban) | 1.05 | 0.93 | 1.19 | 0.455 | **1.37** | 1.11 | 1.68 | 0.003 |
| Keyworker | **1.18** | 1.01 | 1.38 | 0.041 | 1.24 | 0.99 | 1.57 | 0.067 |
| No long-term physical health condition | 1.14 | 0.99 | 1.30 | 0.067 | 1.09 | 0.89 | 1.33 | 0.393 |
| No long-term mental health condition | 1.02 | 0.86 | 1.21 | 0.816 | 1.12 | 0.88 | 1.43 | 0.347 |
| Low confidence in government to handle pandemic | 0.98 | 0.86 | 1.13 | 0.795 | 1.15 | 0.94 | 1.41 | 0.175 |
| Low confidence in health system to handle pandemic | **1.18** | 1.01 | 1.38 | 0.038 | 1.22 | 0.98 | 1.50 | 0.071 |
| Low knowledge of COVID-19 | **1.47** | 1.26 | 1.72 | <0.001 | 1.23 | 0.98 | 1.55 | 0.073 |
| Poor compliance with COVID-19 guidelines | **1.60** | 1.38 | 1.85 | <0.001 | **1.77** | 1.40 | 2.23 | <0.001 |
| Have had COVID-19 | **1.17** | 1.00 | 1.37 | 0.044 | 1.05 | 0.85 | 1.30 | 0.645 |
| No Coronavirus Anxiety Scale symptoms | 1.14 | 0.98 | 1.32 | 0.097 | **1.35** | 1.07 | 1.71 | 0.013 |
| Mistrust of vaccine benefits (ref low) |  |  |  |  |  |  |  |  |
| Intermediate mistrust | **3.36** | 2.87 | 3.93 | <0.001 | **4.98** | 3.90 | 6.37 | <0.001 |
| High mistrust | 1.12 | 0.81 | 1.55 | 0.483 | **4.94** | 3.61 | 6.76 | <0.001 |
| Worries about unforeseen future vaccine effects (ref low) |  |  |  |  |  |  |  |  |
| Intermediate worries | **1.49** | 1.27 | 1.74 | <0.001 | **1.63** | 1.28 | 2.08 | <0.001 |
| High worries | **2.43** | 1.97 | 2.99 | <0.001 | **4.91** | 3.76 | 6.42 | <0.001 |
| Concerns about commercial profiteering (ref low) |  |  |  |  |  |  |  |  |
| Intermediate concerns | **1.95** | 1.69 | 2.26 | <0.001 | **1.73** | 1.34 | 2.24 | <0.001 |
| High level of concern | 0.98 | 0.72 | 1.36 | 0.925 | 1.45 | 0.99 | 2.12 | 0.056 |
| Preference for natural immunity (ref low) |  |  |  |  |  |  |  |  |
| Intermediate preference | **1·63** | 1·42 | 1·87 | <0·001 | **1·77** | 1·41 | 2·22 | <0·001 |
| High preference | **1·50** | 1·13 | 1·99 | 0·005 | **2·51** | 1·78 | 3·53 | <0·001 |
| Did not receive flu vaccine last year | **1·93** | 1·67 | 2·23 | <0·001 | **3·40** | 2·75 | 4·20 | <0·001 |
| Have ever refused a recommended vaccine | **1·44** | 1·19 | 1·75 | <0·001 | **2·54** | 2·00 | 3·23 | <0·001 |
| Constant | 0·02 | 0·01 | 0·03 | <0·001 | 0·00 | 0·00 | 0·00 | <0·001 |

Poor compliance with COVID-19 guidelines and low knowledge about COVID-19 also predicted both vaccine hesitancy and vaccine unwillingness. Further, people who did not receive a flu vaccine last year were twice as likely to be unsure about a COVID-19 vaccine (RRR = 1·93; 95% CI: 1·67 to 2·23) and 3·4 times more likely to have decided against having a COVID-19 vaccine (RRR = 3·40; 95% CI: 2·75 to 4·20).

Demographically, groups at increased risk for uncertainty and unwillingness to vaccinate against COVID-19 were women (uncertain: RRR =1·45; 95% CI: 1·27 to 1·65; unwilling: RRR = 1·52; 95% CI: 1·24 to 1·86), those who were keyworkers (uncertain: RRR = 1·18; 95% CI: 1·01 to 1·38), and people living with children (uncertain: RRR = 1·38; 95% CI: 1·13 to 1·70; unwilling: RRR = 1·60; 95% CI: 1·24 to 2·08).

A socio-economic gradient was also evident for uncertainty and unwillingness to receive the COVID-19 vaccine, with people with lower levels of education increasingly more likely to be unwilling and those with lower incomes more likely to be uncertain. Age was unrelated to uncertainty around the COVID-19 vaccine and only slightly related to unwillingness, with adults over 65 more likely to be willing than younger adults (ages 30-49 and 50-64) to get the COVID-19 vaccine.

Finally, ethnicity, long-term mental and physical health conditions, and low confidence in government to handle the pandemic were unrelated to intentions relating to the COVID-19 vaccine.

## Discussion

This is the first study to comprehensively describe predictors of negative vaccine attitudes and factors influencing uncertainty and unwillingness to vaccinate against COVID-19. Concerningly, groups most at risk of mistrust and misinformation about vaccines are the same groups also at increased risk for illness and death from COVID-19; ethnic minorities and those from socioeconomically deprived backgrounds.^21,22^ The latter, along with women, people with children in the home are also more likely to be uncertain or unwilling to vaccinate against COVID-19 which is consistent with prior work in the US, ^7^ France,^6^ and Australia.^10^ Our findings suggest that the largest behavioural and attitudinal barriers to receiving a COVID-19 vaccine are a general mistrust in the benefits and safety of vaccines and concerns about their unforeseen effects. This echoes some previous work showing that low vaccine confidence and concerns about the novelty and safety of the COVID-19 vaccine are key barriers to vaccine willingness.^7,8,12^ Other substantial behavioural and attitudinal barriers include poor compliance with COVID-19 government guidelines and low knowledge about COVID-19..

Our findings are particularly worrisome given the announcement by the UK government on 5 October 2020 that not only will less than half the population will be able to receive a COVID-19 vaccine, but the vaccine will only be for adults ages 18 and over, certain key workers, the vulnerable, and those over the age of 50.^23^ Our results suggest higher uptake among older adults (65 years plus) compared to young and middle-aged adults (30-49 years and 50-64 years), but no difference in likelihood amongst keyworkers and those with long-term health conditions. As levels of misinformation are no different amongst those with and without long-term health conditions, results could therefore indicate that there will be a demand for the vaccine even amongst people without physical health conditions, which may need to be carefully managed. Potentially more problematic is that individuals of lower socio-economic position are more likely to be undecided or unwilling to be vaccinated, which could exacerbate existing inequalities in exposure to and experience of the virus in the UK.^21,22^

This study also specifically examined factors that predict uncertainty vs unwillingness to be vaccinated. Individuals who are uncertain may be a stronger group for potential interventions.^13^ The uncertain group made up nearly a quarter of our sample (22%) which was a larger proportion than those who were unwilling (14%). This echoes findings from large scale European studies^5^ and in the UK.^8^ Notably, our research suggests that whilst certain factors predict unwillingness but not uncertainty (such as education, age and living in a rural location), it is very difficult to isolate those groups who are merely uncertain. This means that public health campaigns aimed at increasing COVID-19 vaccine uptake should focus on educating both those who are uncertain and those who are unwilling on the safety, efficacy, and side effect profile of vaccines, the importance of complying with social distancing guidelines, and providing clear information on the virus and disease itself.^7,8,10,13^ However, broader public health campaigns to include those who are already willing may also be beneficial in helping them to engage more effectively when they encounter misinformation.^13^

This study has a number of strengths including its large sample size, its longitudinal tracking of participants, and its rich inclusion of measures on psychological and social experiences during COVID-19. However, there are several limitations. The study is not nationally representative, although it does have good stratification across all major socio-demographic groups and analyses were weighted on the basis of population estimates of core demographics. Whilst the recruitment strategy deliberately over-sampled from groups such as ethnic minorities, it is possible that more extreme views on vaccines were not adequately captured. Because we lacked statistical power to look in more detail at sub-groups of different ethnicities, our binary representation likely led to an over-simplification of these diverse categories. Further, this analysis focused on attitudes towards vaccination at the start of the autumn 2020 as the second wave of the virus was beginning in the UK. Future research tracking changing attitudes towards vaccination will be important as this pandemic continues and if and when a vaccination is approved.

Our findings suggest widespread misinformation and anti-vaccine attitudes amongst the general UK public. Many of the specific groups with the most misinformation about vaccines and who are less likely to vaccinate against COVID-19 are also at highest risk for becoming seriously ill with and dying from COVID-19. Despite calculations that more than two-thirds of the public will need to be vaccinated to bring the pandemic under control^1^ and vaccination being central to the UK government’s COVID-19 recovery strategy,^24^ less than half the UK population will be offered a COVID-19 vaccine when it becomes available.^23^ This low distributional goal combined with widespread negative attitudes towards vaccines point to the urgency of developing public health messaging which emphasises vaccine safety. Substantial work has already been undertaken to develop resources for policy makers and other stakeholders to guide effective confidence-building in vaccines are publicly available from the World Health Organization,^25^ Public Health England,^26^ the Centers for Disease Control,^27^, and the European Centre for Disease Prevention and Control.^28^ The research presented here provides a steer as to the demographic groups who most need to be reached if we are to increase vaccine uptake rates at the point a vaccine is available.

## Data Availability

The COVID-19 Social Study documentation and codebook are available for download at https://www.covidsocialstudy.org/. Statistical code is available on request from Elise Paul.

## Contributors

DF designed the study. Data were analysed by EP. EP drafted the manuscript with input from all authors. All authors approved the final version of the manuscript.

## Acknowledgements

The researchers are grateful for the support of a number of organisations with their recruitment efforts including: the UKRI Mental Health Networks, Find Out Now, UCL BioResource, SEO Works, FieldworkHub, and Optimal Workshop.

## Ethics approval and consent to participate

Ethical approval for the COVID-19 Social Study was granted by the UCL Ethics Committee. All participants provided fully informed consent and the study is GDPR compliant.

## Supplementary Material

**Table S1.**
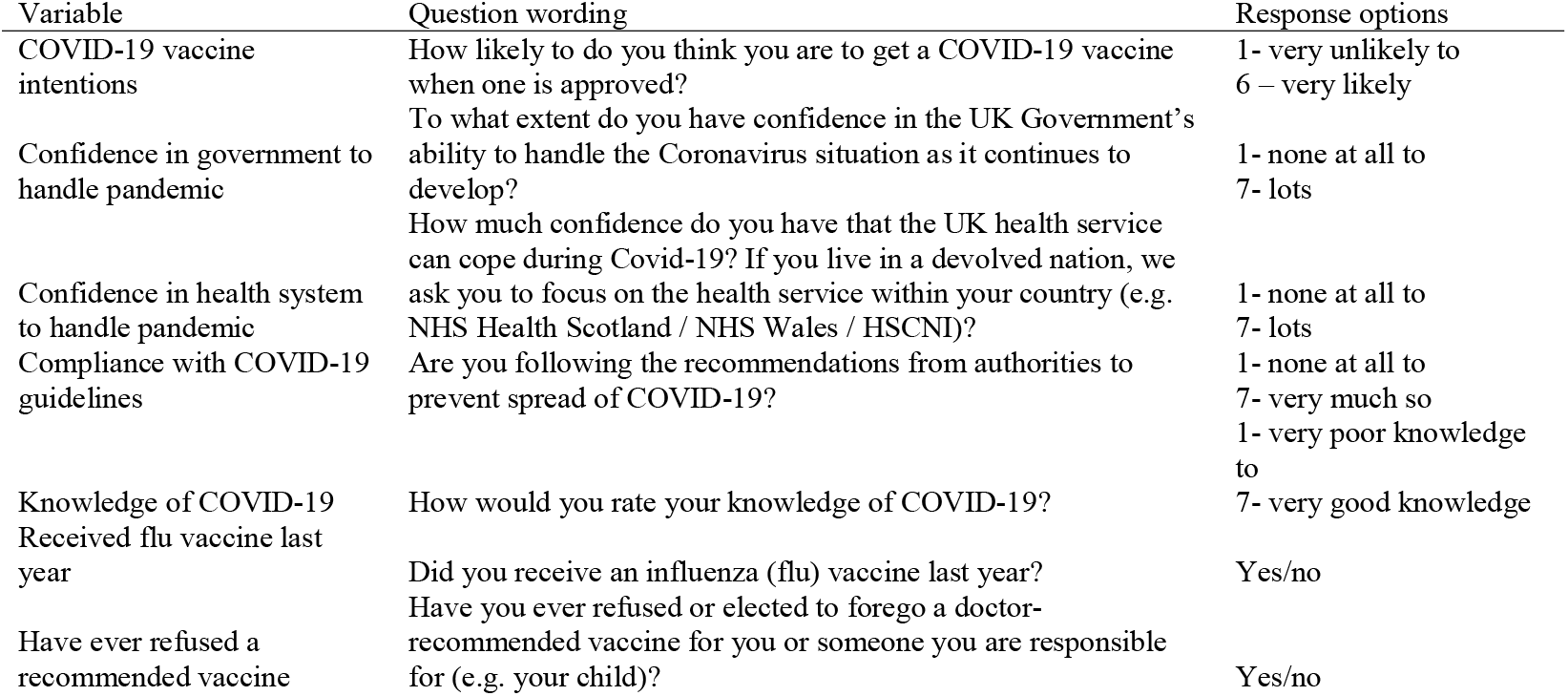
Wording of study-developed items.

**Table S2.** Pattern of missing data in study sample (N = 32,361)

|  | Prop. missing |
| --- | --- |
| Gender | 0 |
| Age | 0 |
| Ethnicity | 0 |
| Education | 0 |
| Income | 9.95% |
| Employment status | 0 |
| Living arrangement | 0 |
| Area of dwelling | 0 |
| Key worker status | 0 |
| Long-term physical health condition | 0 |
| Long-term mental health condition | 0 |
| Confidence in government to handle pandemic | 0.26% |
| Confidence in health system to handle pandemic | 0.25% |
| Knowledge of COVID-19 | 0.25% |
| Compliance with COVID-19 guidelines | 0.25% |
| Have had COVID-19 | 0.10% |
| Coronavirus Anxiety Symptoms scale | 19.66% |
| Prior flu vaccine behaviour | 0.56% |
| Prior vaccine refusal | 0.29% |

**Table S3.**
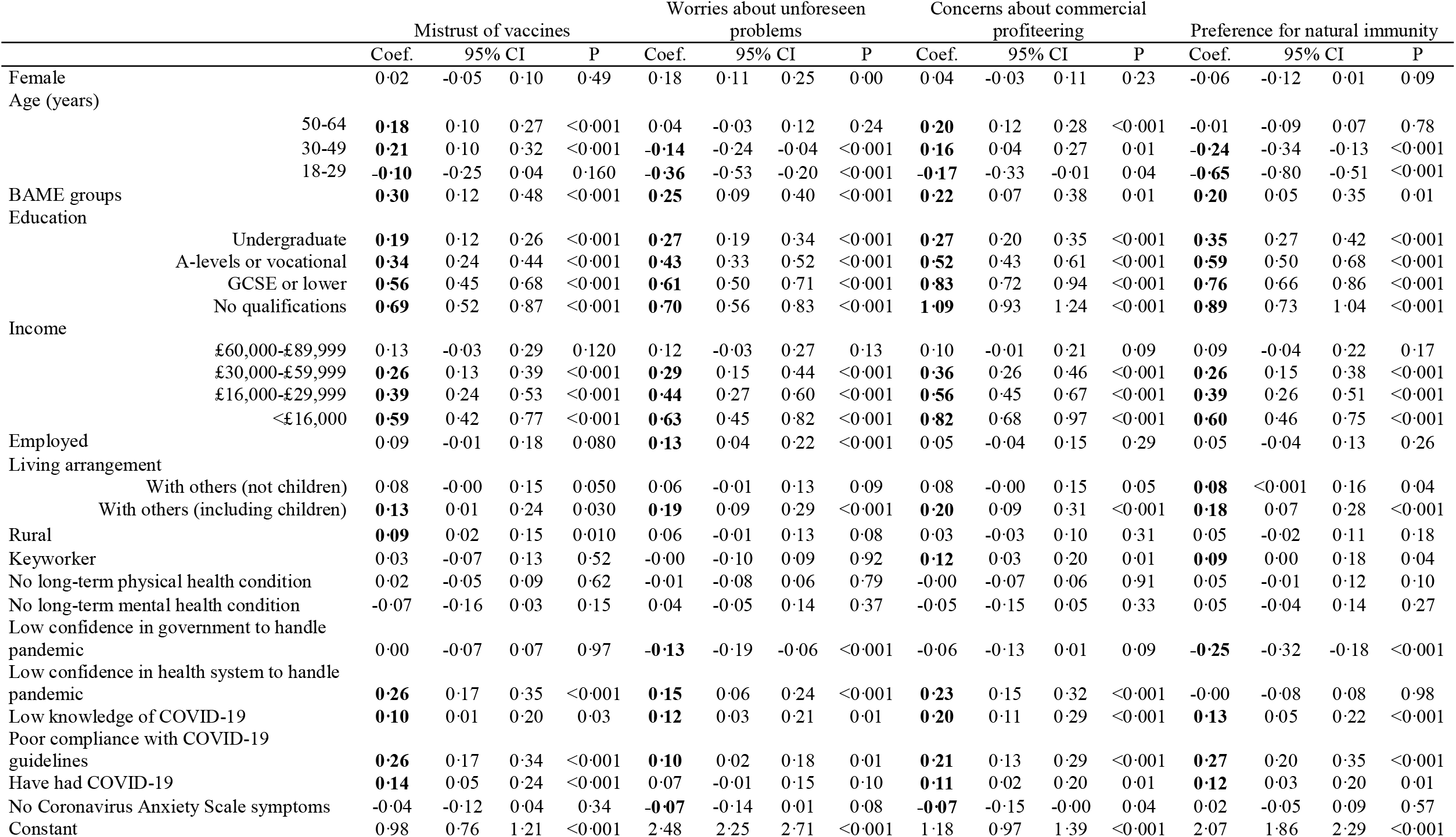
Multivariate linear regression predictors of negative attitudes towards vaccines, complete case analysis (weighted, *N* = 23,164)

|  |  | Mistrust of vaccines |  |  |  | Worries about unforeseen problems |  |  |  | Concerns about commercial profiteering |  |  |  | Preference for natural immunity |  |  |  |
| --- | --- | --- | --- | --- | --- | --- | --- | --- | --- | --- | --- | --- | --- | --- | --- | --- | --- |
|  |  | Coef. | 95% CI |  | P | Coef. | 95% CI |  | P | Coef. | 95% CI |  | P | Coef. | 95% CI |  | P |
| Female |  | 0.02 | -0.05 | 0.10 | 0.49 | 0.18 | 0.11 | 0.25 | 0.00 | 0.04 | -0.03 | 0.11 | 0.23 | -0.06 | -0.12 | 0.01 | 0.09 |
| Age (years) |  |  |  |  |  |  |  |  |  |  |  |  |  |  |  |  |  |
|  | 50-64 | **0.18** | 0.10 | 0.27 | <0.001 | 0.04 | -0.03 | 0.12 | 0.24 | **0.20** | 0.12 | 0.28 | <0.001 | -0.01 | -0.09 | 0.07 | 0.78 |
|  | 30-49 | **0.21** | 0.10 | 0.32 | <0.001 | **-0.14** | -0.24 | -0.04 | <0.001 | **0.16** | 0.04 | 0.27 | 0.01 | **-0.24** | -0.34 | -0.13 | <0.001 |
|  | 18-29 | **-0.10** | -0.25 | 0.04 | 0.160 | **-0.36** | -0.53 | -0.20 | <0.001 | **-0.17** | -0.33 | -0.01 | 0.04 | **-0.65** | -0.80 | -0.51 | <0.001 |
| BAME groups |  | **0.30** | 0.12 | 0.48 | <0.001 | **0.25** | 0.09 | 0.40 | <0.001 | **0.22** | 0.07 | 0.38 | 0.01 | **0.20** | 0.05 | 0.35 | 0.01 |
| Education |  |  |  |  |  |  |  |  |  |  |  |  |  |  |  |  |  |
|  | Undergraduate | **0.19** | 0.12 | 0.26 | <0.001 | **0.27** | 0.19 | 0.34 | <0.001 | **0.27** | 0.20 | 0.35 | <0.001 | **0.35** | 0.27 | 0.42 | <0.001 |
|  | A-levels or vocational | **0.34** | 0.24 | 0.44 | <0.001 | **0.43** | 0.33 | 0.52 | <0.001 | **0.52** | 0.43 | 0.61 | <0.001 | **0.59** | 0.50 | 0.68 | <0.001 |
|  | GCSE or lower | **0.56** | 0.45 | 0.68 | <0.001 | **0.61** | 0.50 | 0.71 | <0.001 | **0.83** | 0.72 | 0.94 | <0.001 | **0.76** | 0.66 | 0.86 | <0.001 |
|  | No qualifications | **0.69** | 0.52 | 0.87 | <0.001 | **0.70** | 0.56 | 0.83 | <0.001 | **1.09** | 0.93 | 1.24 | <0.001 | **0.89** | 0.73 | 1.04 | <0.001 |
| Income |  |  |  |  |  |  |  |  |  |  |  |  |  |  |  |  |  |
|  | £60,000-£89,999 | 0.13 | -0.03 | 0.29 | 0.120 | 0.12 | -0.03 | 0.27 | 0.13 | 0.10 | -0.01 | 0.21 | 0.09 | 0.09 | -0.04 | 0.22 | 0.17 |
|  | £30,000-£59,999 | **0.26** | 0.13 | 0.39 | <0.001 | **0.29** | 0.15 | 0.44 | <0.001 | **0.36** | 0.26 | 0.46 | <0.001 | **0.26** | 0.15 | 0.38 | <0.001 |
|  | £16,000-£29,999 | **0.39** | 0.24 | 0.53 | <0.001 | **0.44** | 0.27 | 0.60 | <0.001 | **0.56** | 0.45 | 0.67 | <0.001 | **0.39** | 0.26 | 0.51 | <0.001 |
|  | <£16,000 | **0.59** | 0.42 | 0.77 | <0.001 | **0.63** | 0.45 | 0.82 | <0.001 | **0.82** | 0.68 | 0.97 | <0.001 | **0.60** | 0.46 | 0.75 | <0.001 |
| Employed |  | 0.09 | -0.01 | 0.18 | 0.080 | **0.13** | 0.04 | 0.22 | <0.001 | 0.05 | -0.04 | 0.15 | 0.29 | 0.05 | -0.04 | 0.13 | 0.26 |
| Living arrangement |  |  |  |  |  |  |  |  |  |  |  |  |  |  |  |  |  |
|  | With others (not children) | 0.08 | -0.00 | 0.15 | 0.050 | 0.06 | -0.01 | 0.13 | 0.09 | 0.08 | -0.00 | 0.15 | 0.05 | **0.08** | <0.001 | 0.16 | 0.04 |
|  | With others (including children) | **0.13** | 0.01 | 0.24 | 0.030 | **0.19** | 0.09 | 0.29 | <0.001 | **0.20** | 0.09 | 0.31 | <0.001 | **0.18** | 0.07 | 0.28 | <0.001 |
| Rural |  | **0.09** | 0.02 | 0.15 | 0.010 | 0.06 | -0.01 | 0.13 | 0.08 | 0.03 | -0.03 | 0.10 | 0.31 | 0.05 | -0.02 | 0.11 | 0.18 |
| Keyworker |  | 0.03 | -0.07 | 0.13 | 0.52 | -0.00 | -0.10 | 0.09 | 0.92 | **0.12** | 0.03 | 0.20 | 0.01 | **0.09** | 0.00 | 0.18 | 0.04 |
| No long-term physical health condition |  | 0.02 | -0.05 | 0.09 | 0.62 | -0.01 | -0.08 | 0.06 | 0.79 | -0.00 | -0.07 | 0.06 | 0.91 | 0.05 | -0.01 | 0.12 | 0.10 |
| No long-term mental health condition |  | -0.07 | -0.16 | 0.03 | 0.15 | 0.04 | -0.05 | 0.14 | 0.37 | -0.05 | -0.15 | 0.05 | 0.33 | 0.05 | -0.04 | 0.14 | 0.27 |
| Low confidence in government to handle pandemic |  | 0.00 | -0.07 | 0.07 | 0.97 | **-0.13** | -0.19 | -0.06 | <0.001 | -0.06 | -0.13 | 0.01 | 0.09 | **-0.25** | -0.32 | -0.18 | <0.001 |
| Low confidence in health system to handle pandemic |  | **0.26** | 0.17 | 0.35 | <0.001 | **0.15** | 0.06 | 0.24 | <0.001 | **0.23** | 0.15 | 0.32 | <0.001 | -0.00 | -0.08 | 0.08 | 0.98 |
| Low knowledge of COVID-19 |  | **0.10** | 0.01 | 0.20 | 0.03 | **0.12** | 0.03 | 0.21 | 0.01 | **0.20** | 0.11 | 0.29 | <0.001 | **0.13** | 0.05 | 0.22 | <0.001 |
| Poor compliance with COVID-19 guidelines |  | **0.26** | 0.17 | 0.34 | <0.001 | **0.10** | 0.02 | 0.18 | 0.01 | **0.21** | 0.13 | 0.29 | <0.001 | **0.27** | 0.20 | 0.35 | <0.001 |
| Have had COVID-19 |  | **0.14** | 0.05 | 0.24 | <0.001 | 0.07 | -0.01 | 0.15 | 0.10 | **0.11** | 0.02 | 0.20 | 0.01 | **0.12** | 0.03 | 0.20 | 0.01 |
| No Coronavirus Anxiety Scale symptoms |  | -0.04 | -0.12 | 0.04 | 0.34 | **-0.07** | -0.14 | 0.01 | 0.08 | **-0.07** | -0.15 | -0.00 | 0.04 | 0.02 | -0.05 | 0.09 | 0.57 |
| Constant |  | 0.98 | 0.76 | 1.21 | <0.001 | 2.48 | 2.25 | 2.71 | <0.001 | 1.18 | 0.97 | 1.39 | <0.001 | 2.07 | 1.86 | 2.29 | <0.001 |

**Table S4.** Predictors of uncertainty and unwillingness to vaccinate against COVID-19 using multivariate multinomial logistic regressions, complete case analysis (weighted, *N* = 23,164)

|  | Undecided COVID-19 |  |  |  | Do not intend COVID-19 |  |  |  |
| --- | --- | --- | --- | --- | --- | --- | --- | --- |
|  | RRR | 95% CI |  | P | RRR | 95% CI |  | P |
| Female (ref male) | **1.43** | 1.24 | 1.65 | <0.001 | **1.69** | 1.38 | 2.08 | <0.001 |
| Age (ref 65+) |  |  |  |  |  |  |  |  |
| 50-64 | 1.15 | 0.94 | 1.40 | 0.173 | **1.32** | 1.01 | 1.73 | 0.042 |
| 30-49 | 1.24 | 0.97 | 1.59 | 0.085 | **1.49** | 1.08 | 2.06 | 0.016 |
| 18-29 | 1.41 | 1.00 | 1.98 | 0.051 | 1.30 | 0.81 | 2.07 | 0.275 |
| Ethnicity (ref White) |  |  |  |  |  |  |  |  |
| BAME groups | 1.09 | 0.82 | 1.45 | 0.538 | 1.32 | 0.88 | 1.98 | 0.177 |
| Education (ref postgraduate) |  |  |  |  |  |  |  |  |
| Undergraduate | 0.97 | 0.82 | 1.14 | 0.695 | **1.37** | 1.06 | 1.79 | 0.017 |
| A-levels or vocational | 1.09 | 0.89 | 1.33 | 0.428 | **1.82** | 1.35 | 2.45 | <0.001 |
| GCSE or lower | 1.20 | 0.97 | 1.48 | 0.101 | **2.36** | 1.70 | 3.28 | <0.001 |
| No qualifications | 1.16 | 0.84 | 1.62 | 0.369 | **2.84** | 1.79 | 4.49 | <0.001 |
| Income (ref >£90,000) |  |  |  |  |  |  |  |  |
| £60,000-£89,999 | 1.23 | 0.91 | 1.65 | 0.175 | 0.94 | 0.59 | 1.50 | 0.808 |
| £30,000-£59,999 | **1.48** | 1.17 | 1.89 | 0.001 | 1.20 | 0.80 | 1.81 | 0.381 |
| £16,000-£29,999 | **1.59** | 1.22 | 2.07 | 0.001 | 1.38 | 0.89 | 2.13 | 0.146 |
| <£16,000 | **2.23** | 1.67 | 2.99 | <0.001 | **1.91** | 1.18 | 3.08 | 0.008 |
| Employed | **1.26** | 1.04 | 1.51 | 0.017 | 1.12 | 0.85 | 1.46 | 0.422 |
| Living arrangement (ref live alone) |  |  |  |  |  |  |  |  |
| With others (not children) | 1.07 | 0.90 | 1.27 | 0.432 | 1.07 | 0.84 | 1.36 | 0.572 |
| With others (including children) | **1.33** | 1.06 | 1.67 | 0.014 | **1.51** | 1.12 | 2.04 | 0.007 |
| Rural | 1.12 | 0.97 | 1.30 | 0.115 | **1.38** | 1.10 | 1.71 | 0.004 |
| Key worker | 1.08 | 0.90 | 1.30 | 0.414 | 1.07 | 0.82 | 1.40 | 0.609 |
| No long-term physical health condition | **1.19** | 1.03 | 1.38 | 0.021 | 1.11 | 0.90 | 1.37 | 0.316 |
| No long-term mental health condition | 0.99 | 0.82 | 1.19 | 0.884 | 1.11 | 0.84 | 1.45 | 0.469 |
| Low confidence in government to handle pandemic | 0.97 | 0.83 | 1.13 | 0.686 | 1.15 | 0.93 | 1.43 | 0.202 |
| Low confidence in health system to handle pandemic | **1.29** | 1.09 | 1.53 | 0.004 | **1.52** | 1.20 | 1.93 | <0.001 |
| Low knowledge of COVID-19 | **1.33** | 1.12 | 1.59 | 0.001 | 1.22 | 0.95 | 1.57 | 0.123 |
| Poor compliance with COVID-19 guidelines | **1.50** | 1.27 | 1.76 | <0.001 | **2.13** | 1.70 | 2.67 | <0.001 |
| Have had COVID-19 | 1.08 | 0.90 | 1.29 | 0.387 | 1.07 | 0.84 | 1.36 | 0.593 |
| No Coronavirus Anxiety Scale symptoms | **1.21** | 1.04 | 1.40 | 0.011 | **1.38** | 1.10 | 1.74 | 0.005 |
| Mistrust of vaccine benefits (ref low) |  |  |  |  |  |  |  |  |
| Intermediate mistrust | **3.70** | 3.08 | 4.46 | <0.001 | **5.58** | 4.31 | 7.22 | <0.001 |
| High mistrust | 1.22 | 0.85 | 1.76 | 0.286 | **5.13** | 3.54 | 7.44 | <0.001 |
| Worries about unforeseen future vaccine effects (ref low) |  |  |  |  |  |  |  |  |
| Intermediate worries | **1.59** | 1.34 | 1.88 | <0.001 | **1.44** | 1.08 | 1.90 | 0.012 |
| High worries | **2.77** | 2.18 | 3.51 | <0.001 | **5.61** | 4.08 | 7.72 | <0.001 |
| Concerns about commercial profiteering (ref low) |  |  |  |  |  |  |  |  |
| Intermediate concerns | **1.85** | 1.56 | 2.18 | <0.001 | **1.50** | 1.16 | 1.95 | 0.002 |
| High level of concern | 0.76 | 0.53 | 1.10 | 0.145 | 1.10 | 0.72 | 1.68 | 0.654 |
| Preference for natural immunity (ref low) | **1.67** | 1.43 | 1.96 | <0.001 | **1.88** | 1.46 | 2.43 | <0.001 |
| Intermediate preference |  |  |  |  |  |  |  |  |
| High preference | **1.82** | 1.33 | 2.48 | <0.001 | **3.27** | 2.24 | 4.77 | <0.001 |
| Did not receive flu vaccine last year | **1.92** | 1.65 | 2.23 | <0.001 | **3.47** | 2.82 | 4.27 | <0.001 |
| Have ever refused a recommended vaccine | **1.39** | 1.12 | 1.73 | 0.003 | **2.40** | 1.85 | 3.13 | <0.001 |
| Constant | 0.02 | 0.01 | 0.02 | <0.001 | 0.00 | 0.00 | 0.00 | <0.001 |

